# Immunogenicity and reactogenicity against the SARS-CoV-2 variants following heterologous primary series involving CoronaVac and ChAdOx1 and BNT162b2 plus heterologous BNT162b2 booster vaccination: An open-label randomized study in healthy Thai adults

**DOI:** 10.1101/2022.03.03.22271601

**Authors:** Suvimol Niyomnaitham, Zheng Quan Toh, Patimaporn Wongprompitak, Laddawan Jansarikit, Kanjana Srisutthisamphan, Sompong Sapsutthipas, Yuparat Jantraphakorn, Natthakarn Mingngamsup, Paul V Licciardi, Kulkanya Chokephaibulkit

## Abstract

We evaluated the immunogenicity and reactogenicity of heterologous COVID-19 primary series vaccination schedules. Participants were randomized to one of seven groups that received two-dose homologous BNT162b2 or heterologous combinations of CoronaVac, ChAdOx1 and BNT162b2, with 4 weeks interval. Of 210 participants, median age was 38 (19-60) years, 51% were female. The groups that received BNT162b2 as second dose induced the highest virus-specific IgG response against the ancestral strain [BNT162b2: geometric mean concentration (GMC) 2133-2249, 95%CI 1558 to 3055; ChAdOx1: 851-1201, 95%CI 649 to 1522; CoronaVac: 137-225, 95%CI 103-286 BAU/mL], neutralising antibodies (NAb) against Beta and Delta, and interferon gamma response. All groups induced low to negligible NAb against Omicron. A BNT162b2 booster (3^rd^ dose) following heterologous CoronaVac and ChAdOx1 regimens induced >140-fold increase in NAb titres against Omicron. Our findings indicate that heterologous regimens using BNT162b2 as the second dose may be considered an alternative schedule to maximize immune response.

## Introduction

As of 22^nd^ of Feb 2022, the severe acute respiratory syndrome coronavirus-2 (SARS-CoV-2) has infected more than 400 million people and caused more than 5 million deaths globally^1^. Vaccination against COVID-19 has been crucial for controlling the pandemic, with 9 COVID-19 vaccines receiving World Health Organization (WHO) Emergency Use listing (EUL) to date. The ChAdOx1 (a chimpanzee adenovirus-vectored vaccine expressing the SARS-CoV-2 spike protein, Oxford University-AstraZeneca), BNT162b2 (SARS-CoV-2 spike protein mRNA vaccine, Pfizer-BioNTech, US-Germany) and CoronaVac (an inactivated whole-virion SARS-CoV-2 vaccine, Sinovac Life Science, China) are the most widely used COVID-19 vaccines globally; ChAdOx1 and CoronaVac are used in many low- and middle-income countries^2,3^. All three vaccines given as 2-dose primary schedule were demonstrated to be safe and effective in preventing symptomatic COVID-19 and severe disease caused by the ancestral SARS-CoV-2 Wuhan strain as well as Alpha, Beta and Delta variants^4^. The SARS-CoV-2 Omicron variant identified in late 2021 was found to harbour 36 mutations in the spike protein which enable the virus to evade immunity induced by infection or vaccination. Many studies suggested that a booster dose is needed to protect against Omicron^5–8^.

Although the level of neutralising antibodies to protect against infection or severe disease has not been identified, it is clear that neutralising antibodies correlate with vaccine effectiveness and are believed to be the primary mechanism of protection against SARS-CoV-2 infection^9^. Other aspects of the immune response, in particular cellular immunity, are also likely to be important in protection particularly against severe disease and are important to measure following COVID-19 vaccination.

The limited supply of COVID-19 vaccines globally has resulted in less than 10% of the eligible population in low-income countries being fully vaccinated^10^. Heterologous COVID-19 vaccination or mix-and-match COVID-19 vaccine schedules would alleviate the vaccine supply issues and allow more flexible COVID-19 vaccination in these settings. However, such strategies need to be demonstrated to be equally safe and immunogenic compared with current homologous COVID-19 vaccination. While some studies that reported similar or higher immunogenicity following heterologous primary vaccination involving the mRNA vaccines (BNT162b2 and mRNA-1273: SARS-CoV-2 spike protein mRNA vaccine, Moderna, US) and ChAdOx1^11^ compared to homologous vaccination^12^, limited data is available for heterologous primary vaccination involving inactivated vaccines such as CoronaVac.

In Thailand, due to the lack of access to mRNA vaccine in the early phase of the pandemic, and following early reports of better immunogenicity in heterologous primary series than homologous primary series, a CoronaVac-prime and ChAdOx1-boost schedule has been implemented across the country^13^. However, limited data exists for how these vaccine schedules, as well as following an additional dose of vaccine might protect against the Delta and Omicron variants. We evaluated the reactogenicity and immunogenicity of heterologous COVID-19 primary vaccine schedules involving BNT162b2, ChAdOx1 or CoronaVac, as well as BNT162b2 boosting in participants who received ChAdOx1 and CoronaVac as primary series.

## Results

### Participant’s baseline characteristics

Between January 2021-June 2021, a total of 220 participants were screened and 210 were enrolled. Participants were assigned to one of the seven groups (30 per group) of either two-dose homologous BNT162b2 or heterologous combinations of CoronaVac, ChAdOx1 and BNT162b2 (CoronaVac-ChAdOx1, CoronaVac-BNT162b2, ChAdOx1-CoronaVac, ChAdOx1-BNT162b2, BNT162b2-CoronaVac, BNT162b2-ChAdOx1). Among the participants, eight participants were infected with SARS-COV-2 at baseline, defined by positive anti-nucleoprotein antibody (anti-NP IgG) and were excluded from the analysis, while one participant was lost to follow up (Figure 1). The overall median (range) age of study participants were 38 (19-60) years old, and 108/210 (51.43%) were female. The baseline characteristics were similar across the seven groups (Table 1).

**Table 1.** Baseline characteristics of study participants.

| The groups by vaccine used as the first and second dose |  |  |  |  |  |  |  |
| --- | --- | --- | --- | --- | --- | --- | --- |
|  | CoronaVac<br>-ChAdOx1 | CoronaVac<br>-BNT162b2 | ChAdOx1 -<br>CoronaVac | ChAdOx1-BNT162b2 | BNT162b2 -<br>CoronaVac | BNT162b2 -<br>ChAdOx1 | BNT162b2 -<br>BNT162b2 |
| Numbers of<br>enrolled<br>participants | n=30 | n=30 | n=30 | n=30 | n=30 | n=30 | n=30 |
| Age (years),<br>median,<br>IQR | 39<br>(33, 46) | 30.5<br>(24, 38) | 40.5<br>(36, 48) | 39<br>(29, 43) | 34.5<br>(25, 47) | 32.5<br>(26, 41) | 36.5<br>(32, 43) |
| Female, n<br>(%) | 15 (50) | 18 (60) | 14 (46.67) | 19 (63.33) | 13 (43.3) | 16 (53.3) | 13 (43.3) |
| BMI,<br>median<br>(IQR) | 25.25<br>(22.70,<br>28.10) | 23.40<br>(21.00,<br>26.50) | 23.5<br>(21.30,<br>26.00) | 20.95<br>(19.60,<br>24.50) | 24.15<br>(20.10,<br>27.70) | 24.35<br>(20.80,<br>28.70) | 24.30<br>(21.40,<br>26.10) |
| Hypertensio<br>n, n (%) | 1 (3.33) | 0 | 4 (13.33) | 1 (3.33) | 0 | 2 (6.67) | 0 |
| Dyslipidemi<br>a, n (%) | 0 | 0 | 2 (6.67) | 0 | 0 | 2 (6.67) | 0 |
| Diabetes, n<br>(%) | 0 | 0 | 1 (3.33) | 0 | 0 | 0 | 0 |

**Fig. 1.**
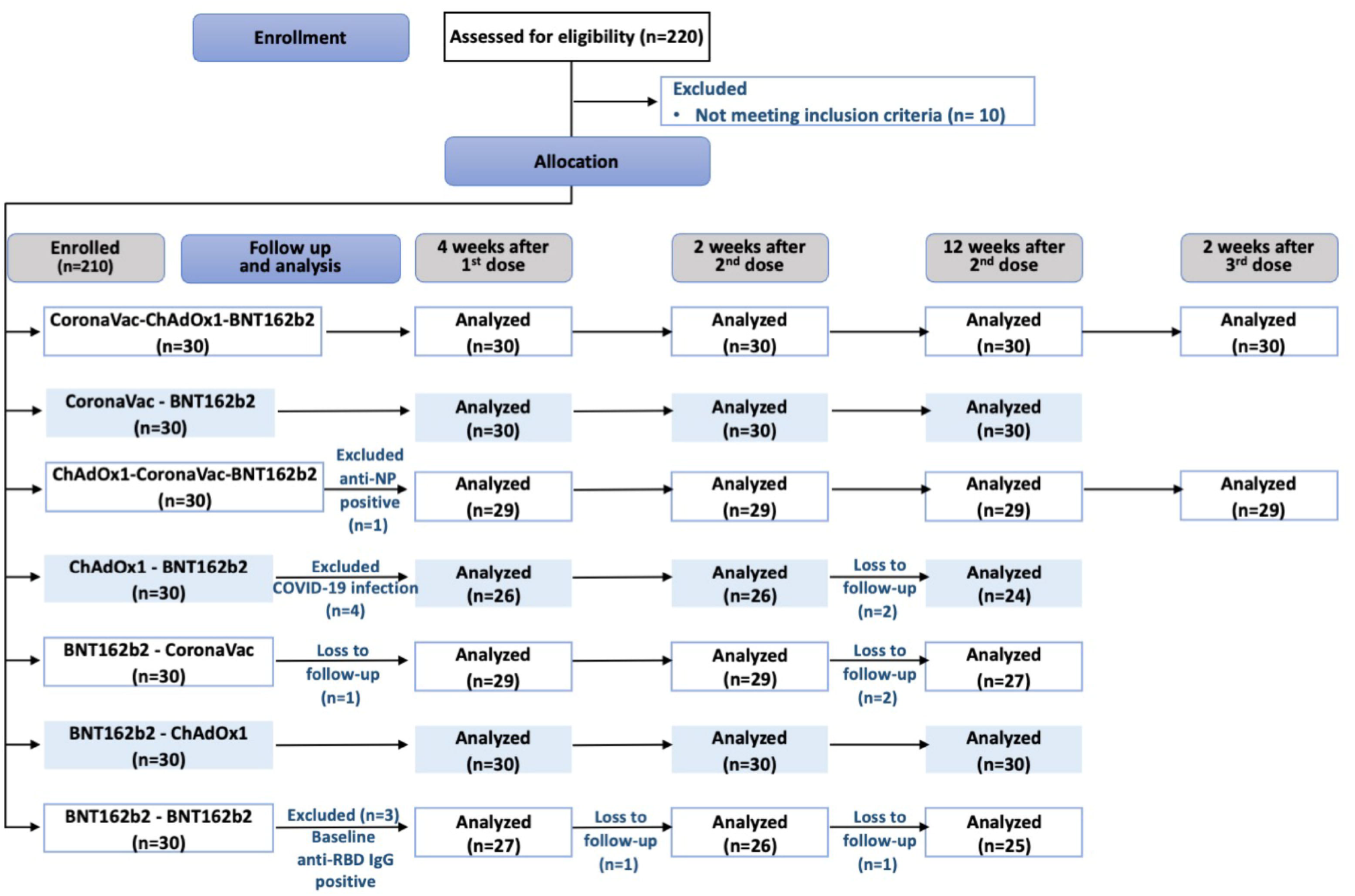
Consort flow diagram describing the allocation and number of analyzed participants in each study group.

### SARS-CoV-2 IgG response against ancestral virus following heterologous primary series

All participants from each group seroconverted at 4 weeks after the first dose. At 2 weeks after the second dose, the virus-specific IgG levels were highest among the groups who received BNT162b2 as second dose (Figure 2A): CoronaVac-BNT162b2 (2181.8 BAU/mL, 95%CI 1558.2 to 3055.1) and ChAdOx1-BNT162b2 groups (2132.7 BAU/mL, 95%CI 1696.1 to 2,681.7); both groups have similar IgG levels compared with the homologous BNT162b2-BNT162b2 group (2248.8 BAU/mL, 95%CI 1691.3 to 2,990.0). These levels were significantly higher compared with the groups who received ChAdOx1 or CoronaVac as second dose (CoronaVac-ChAdOx1: 851.4 BAU/mL, 95%CI 649.5 to 1116.1; BNT162b2-ChAdOx1: 1201.2 BAU/mL, 95%CI 947.9 to 1522.1; ChAdOx1-CoronaVac: 137.04 BAU/mL, 95%CI 103.6 to 186.4; BNT162b2-CoronaVac: 225.2 BAU/mL, 95%CI 177.1 to 286.4) (Figure 2 and Supplementary Table 1). Based on our previous data on homologous ChAdOx1 or CoronaVac primary series at 2 weeks after the second dose^14^ heterologous schedule with ChAdOx1 as second dose (CoronaVac-ChAdOx1or BNT162b2-ChAdOx1) induced significantly higher IgG than the homologous ChAdOx1 or CoronaVac schedules (*p*<0.0001). In contrast, similar or lower IgG levels were observed with heterologous schedule with CoronaVac as second dose (ChAdOx1-CoronaVac or BNT162b2-CoronaVac) (Figure 2 and Supplementary Table 1). The geometric mean ratio (GMR) from dose 1 to dose 2 were highest in those that received CoronaVac as first dose and received either ChAdOx1 (55-fold) or BNT162b2 (110-fold) as second dose. The GMR ratio between dose 1 and 2 in groups that received CoronaVac as the second dose (ChAdOx1-CoronaVac and BNT162b2-CoronaVac) was the lowest (approximately 1.6-fold) (Supplementary Table 1).

**Fig. 2.**
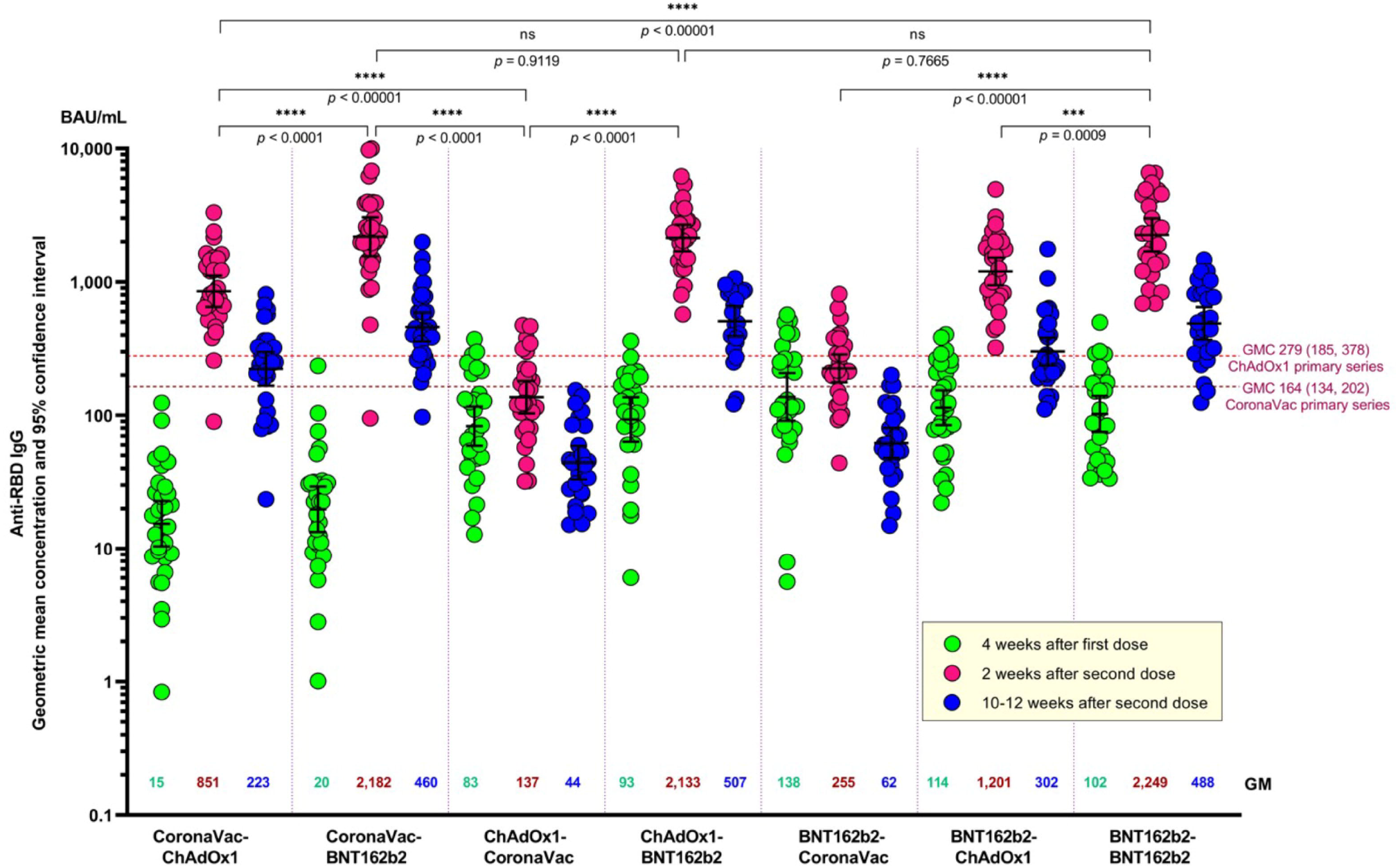
Anti-SARS-CoV-2 receptor binding domain (RBD) IgG at 4 weeks after the first dose, and 2 and 10-12 weeks after the second dose. Numbers in the graph represent geometric mean concentration (GMC) and the error bars represent 95% confidence interval. Dotted lines represent the GMC (95% CI) of anti-SARS-CoV-2 RBD IgG at 2 weeks after the second dose of homologous primary series of CoronaVac and ChAdOx1 reported by our study group using the same laboratory method and facility as this study^4^. Unpaired t-test were used to compare IgG GMC between groups at two weeks after second dose vaccination.

At 10-12 weeks post-second dose, there was around 3- to 4-fold decrease in IgG levels across all groups (Figure 2 and Supplementary Table 1). The groups that received CoronaVac as the second dose had IgG levels significantly lower than the levels found at 4 weeks post first dose (p<0.0001).

### Neutralizing antibody responses against SARS-CoV-2 variants (Delta, Beta and Omicron) following heterologous primary series

Neutralizing antibodies (PRNT_50_) against the Delta and Beta SARS-CoV-2 variants for each group were measured at 2 weeks after the second dose using plaque-reduction neutralization test (Figure 3). Similar to the IgG response against the ancestral Wuhan strain, the groups whom were given BNT162b2 as second dose had significantly higher PRNT_50_ against Delta and Beta than the groups that received ChAdOx1 or CoronaVac as the second dose (PRNT_50_ geometric mean titres (GMT) for Delta: BNT162b2 groups 195.12-196.97 (95%CI 126.9 to 305.7), ChAdOx1 groups 78.65-112.36 (95%CI 57.1 to 147.1), CoronaVac groups 20.36-22.69 (95%CI 14.2 to 32.2), p<0.001; PRNT_50_ for Beta: BNT162b2 groups 43.28-62.36 (95%CI 28.9 to 101.6), ChAdOx1 groups 20.43-30.56 (95%CI 14.4 to 50.4), CoronaVac groups, 8.18-9.01 (95%CI 6.5 to 11.2), p<0.001) (Figure 3A and Supplementary Table 1). The PRNT_50_ in the heterologous groups that received BNT162b2 as second dose were similar to those in the homologous BNT162b2 group. For all groups, PRNT_50_ against the Beta variant were reduced by 2 to 5-fold compared to the Delta variant (Supplementary Table 1).

**Fig. 3.**
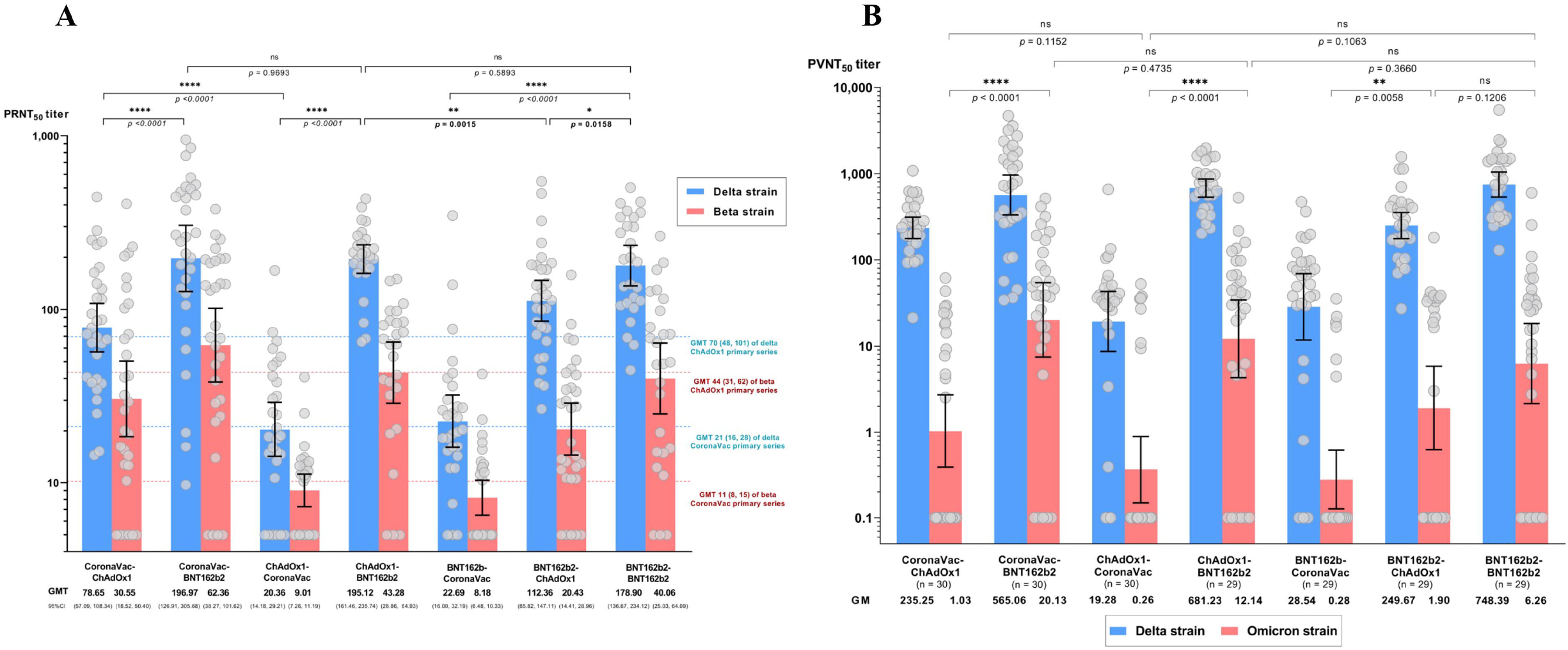
Neutralizing antibody titers against SARS-CoV-2 variants at two weeks after second dose. (A) Plaque reduction neutralization titers (PRNT_50_) against Delta (blue) and Beta (red) variants. The dot line referred to the geometric mean tires at 2 weeks after the second dose of homologous CoronaVac or ChAdOx1 reported by our group using the same laboratory methods and facility with this study^4^. (B) Pseudovirus-based neutralizing antibody titers (PVNT_50_) against Delta (blue) and Omicron variants (red). Numbers on the x-axis represent geometric mean titer (GMT) and error bars represent 95% confidence interval (CI). Unpaired t-test were used to compare PRNT_50_ between each group at two weeks after second dose.

Due to the unavailability of live-virus assay against Omicron, we used pseudovirus-based neutralization assay to evaluate neutralizing antibody activity against Omicron and Delta to compare the two variants. There was a strong correlation between the PVNT_50_ and PRNT_50_ titres against Delta (r=0.79) (Supplementary Figure 1). At 2 weeks after the second dose, the seropositivity rate (defined as >1:10 PVNT_50_) against Omicron among groups who received BNT162b2, ChAdOx1, and CoronaVac as second dose were 80% (45/56), 50% (30/60), and 21% (12/58), respectively. as the seropositivity rate was 62% (18/29) for the homologous 2-dose BNT162b2 group. Overall, PVNT_50_ against Omicron was low across the groups and were 28- to 229-fold lower than Delta, depending on the vaccine schedules (Figure 3B and Supplementary Table 1). The PVNT_50_ against both Delta and Omicron were significantly lower among groups who received CoronaVac as second dose compared to the other groups

### Interferon gamma responses following heterologous primary series

The interferon gamma response was measured using the interferon gamma release assay (IGRA) as a surrogate for SARS-CoV-2-specific T cell responses. At 2 weeks after the second dose, groups who received BNT162b2 as second dose (>80% of participants, including homologous BNT162b2 group) had the highest IGRA positivity rate, followed by the groups who received ChAdOx1 (66-73%) and CoronaVac (55-59%) (Figure 4 and Supplementary Table 2). The IGRA levels are shown in Supplementary Figure 2.

**Fig. 4.**
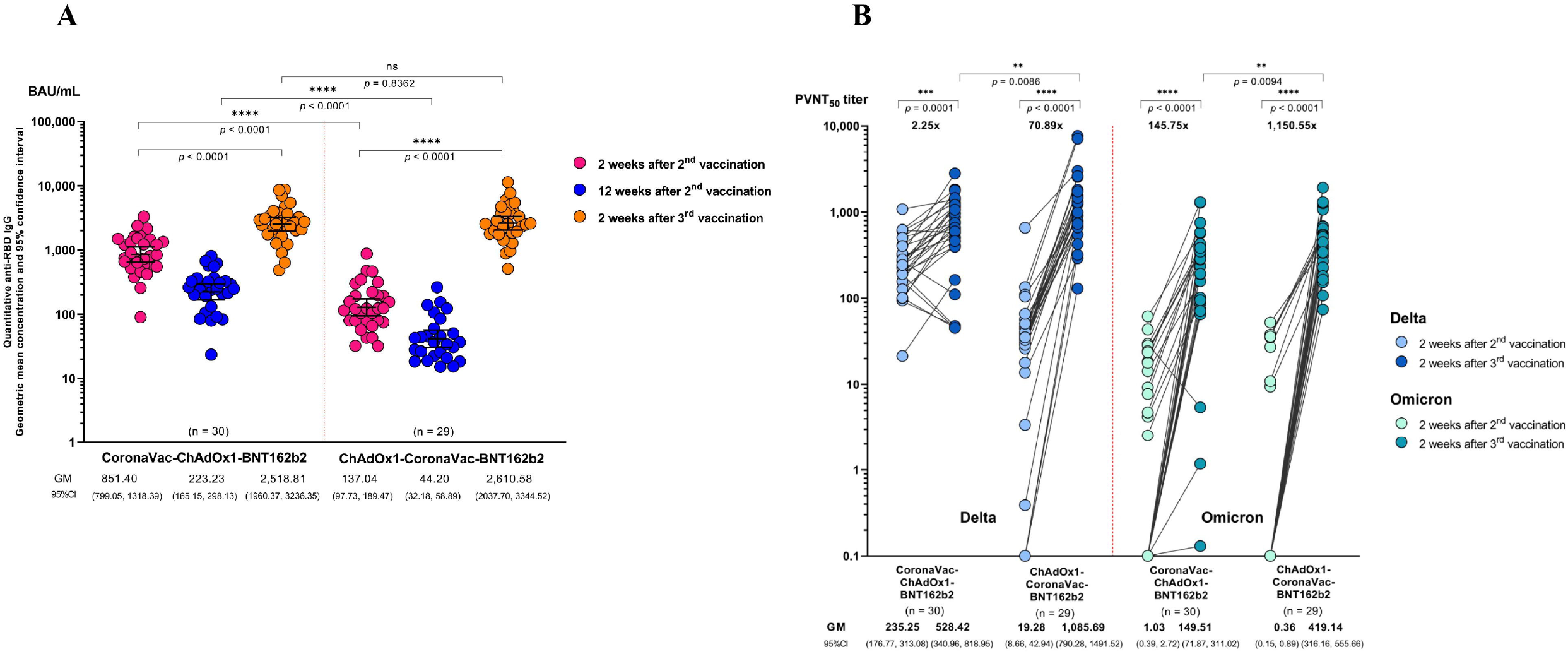
Humoral immune response following a BNT162b2 booster (3rd dose) vaccination in the groups that received heterologous CoronaVac and ChAdOx1 in the primary series. (A) anti-SARS-CoV-2 receptor binding domain (RBD) IgG kinetics at two weeks after second dose (pink), 12 weeks after second dose (blue), and two weeks after the third booster dose (orange). (B) Pseudovirus-based neutralizing antibody titres (PVNT50) against Delta (blue) and Omicron (green) variants following a BNT162b2 booster (third dose) at two weeks after second dose and two weeks after the third booster dose. Numbers on the x-axis represent geometric mean titer. The fold change and significance p-value comparing two weeks after the third booster and two weeks after second dose.

### Immunogenicity of BNT162b2 booster (3^rd^ dose) vaccination in the groups that received heterologous CoronaVac and ChAdOx1 in the primary series

Based on the relatively lower immunogenicity observed in the groups of CoronaVac-ChAdOx1 and ChAdOx1-CoronaVac, and the relevance of these schedules for Thailand and other LMICs, we investigated the immunogenicity of a BNT162b2 booster given at 10-12 weeks after the second dose. At two weeks after the booster dose, the virus-specific IgG levels against the ancestral strain were similar between the CoronaVac-ChAdOx1 and ChAdOx1-CoronaVac groups (2518.8, 95%CI: 1960.4, 3236.4 BAU/mL and 2610.6, 95%CI 2037.7-3344.5 BAU/mL, respectively, *p*=0.84) (Figure 4A and Supplementary Table 1). However, the GMR between two weeks post 3^rd^ and two weeks post 2^nd^ dose were 2.5 and 18.9 for CoronaVac-ChAdOx1-BNT162b2 and ChAdOx1-CoronaVac-BNT162b2 groups, respectively (Supplementary Table 1).

Consistent with this observation, two weeks after a BNT162b2 booster, neutralizing antibodies against Omicron were 145.8- and 1150.6-fold higher than two weeks after the second dose for CoronaVac-ChAdOx1-BNT162b2 and ChAdOx1-CoronaVac-BNT162b2 groups, respectively (Figure 4B and Supplementary Table 1). Notably, the ChAdOx1-CoronaVac-BNT162b2 group had higher neutralising antibodies against Delta and Omicron than CoronaVac-ChAdOx1-BNT162b2 group (Supplementary Table 1). Neutralising antibody titres against Omicron were around 2-3-fold lower than against Delta.

### Reactogenicity of second dose and third dose following heterologous vaccination

The overall adverse events (AE) reported for all groups following the second dose were mild to moderate, with no serious AE reported (Figure 5). Systemic reactions were most frequent among those who received CoronaVac-ChAdOx1 (97%, 29/30), while similar local reactions were observed between those who received BNT162b2 as the second dose, as well as those who received CoronaVac-ChAdOx1 schedule (87%-93%). Those who received CoronaVac as the second dose had significantly lower local reactions (45-53%) or systemic reactions (48-67%) compared to the other groups. The most frequently reported systemic reactions among all the study groups were fatigue, myalgia and headaches. The AE reported after the BNT162b2 booster (third) dose was similar to that reported after the BNT162b2 second dose.

**Fig. 5.**
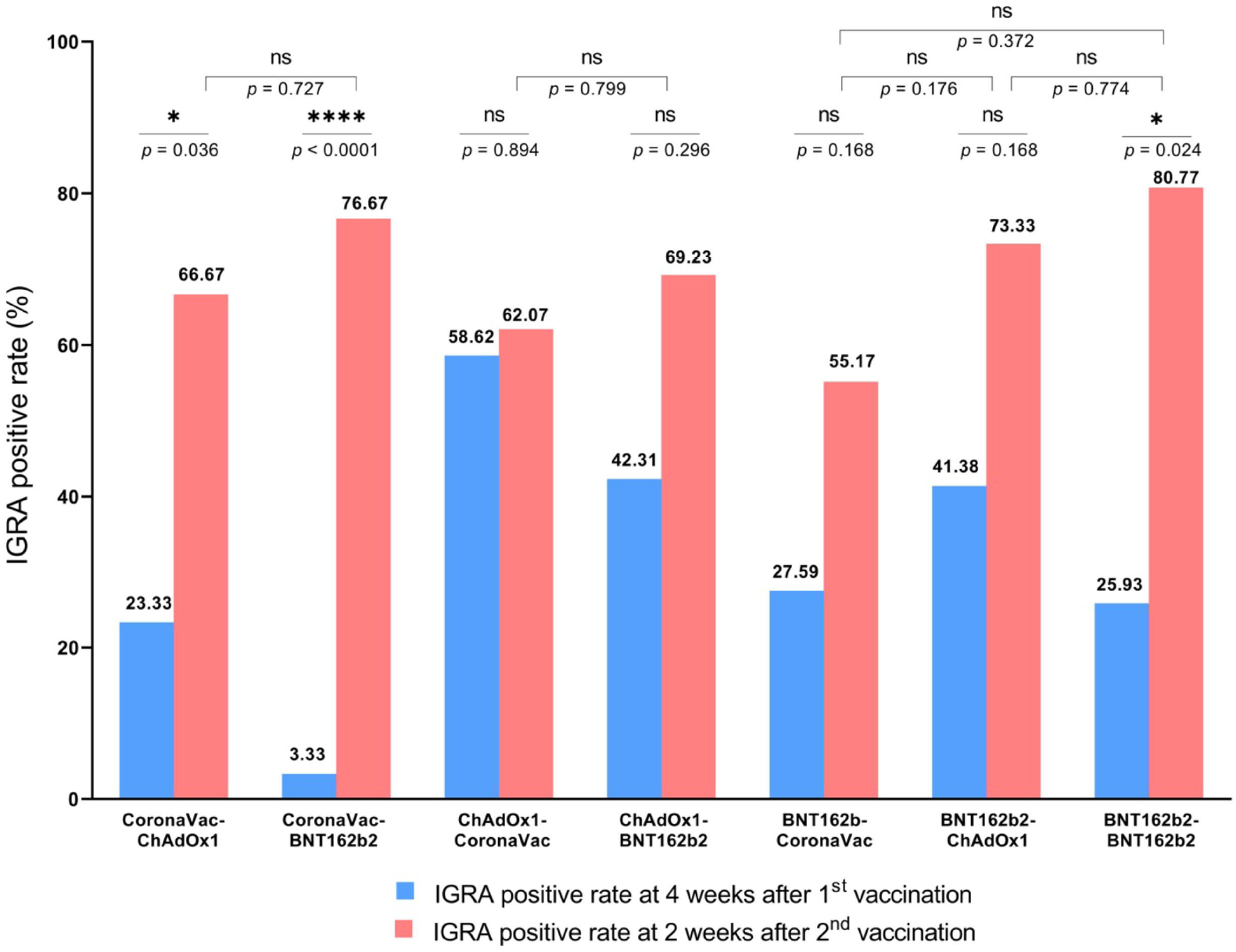
Proportion of interferon-gamma release assay (IGRA)-positivity rate at four weeks after the first dose and two weeks after the second dose. IGRA-positive is defined as positive response to either antigen 1 (Ag1) or antigen 2 (Ag2) of the assay. The proportions of IGRA positive following the first dose vaccination in the groups that received the same vaccine were not significantly different.

**Fig. 6.**
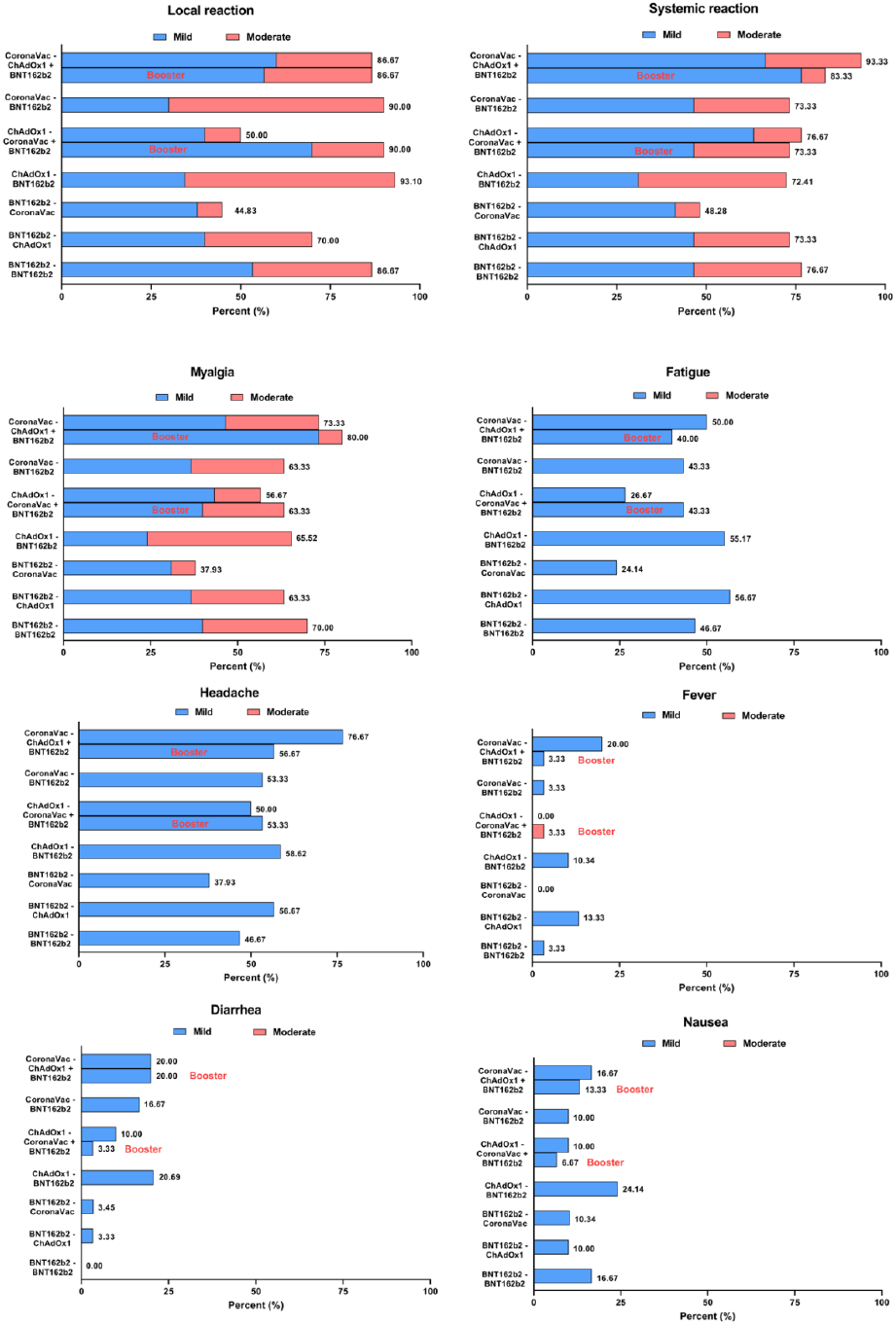
Adverse events following second dose and third dose. Stacked bars represents the proportion of participants who reported mild and moderate adverse events.

## Discussion

Heterologous vaccination may improve the immunogenicity and flexibility of vaccine schedules, particularly when the supply of a vaccine is limited. Our findings showed that heterologous schedules involving BNT162b2, ChAdOx1 and CoronaVac are safe, but their immunogenicity varied depending on the type of vaccine given as the second dose. Of note, using BNT162b2 as the second dose, regardless of the vaccine type given as first dose, induced the highest humoral and cellular immune responses, and is equivalent to homologous BNT162b2 schedules. These schedules however were poorly immunogenic against Omicron. An additional dose BNT162b2 was able to boost the neutralizing antibody response against Omicron in participants who received heterologous CoronaVac-ChAdOx1 or ChAdOx1-CoronaVac. Taken together, heterologous prime-boost schedules using BNT162b2 as the second dose may improve the immunogenicity of homologous CoronaVac and ChAdOx1 primary series, but unlikely to protect against Omicron for which a third booster BNT162b2 is needed.

Our findings on heterologous schedules involving ChAdOx1 and BNT162b2 supports earlier findings from studies predominately from Europe where priming of ChAdOx1 and boosting with BNT162b2 as the second dose induced higher humoral and cell-mediated immune responses than homologous ChAdOx1 primary series^11,12,15–17^, and to some extent higher cell-mediated immune responses than homologous BNT162b2 primary series. Some studies have found better neutralising capacity against the SARS-CoV-2 variants and immune memory responses following heterologous ChAdOx1–BNT162b2 schedule compared to homologous ChAdOx1 or BNT162b2^18^. Whether other combinations of heterologous COVID-19 vaccination, including longer intervals between different vaccines can improve protection against SARS-CoV-2 variants is unknown. The heterologous schedules in this study were given 4 weeks apart and are similar to the intervals for homologous BNT162b2 or CoronaVac vaccination. This interval may provide earlier protection against SARS-CoV-2 infection unlike homologous ChAdOx1 vaccination where the intervals are generally recommended between 8-12 weeks to yield a better protection. This is particularly relevant for protection against the Delta variant, where two doses of the vaccines are needed^19^.

While there have been data on heterologous schedules involving ChAdOx1 and BNT162b2, to our knowledge, only one study has evaluated heterologous primary series involving inactivated vaccine such as CoronaVac^13^. The study found higher immunogenicity of CoronaVac-ChAdOx1 schedule than either homologous CoronaVac or ChAdOx1 vaccination^13^. Consistent with this finding, we found that CoronaVac given as the first priming dose induced sufficient immune memory, which when boosted with either ChAdOx1 and BNT162b2 as the second dose induced high levels of humoral and cellular immunity. However, CoronaVac given as the second dose following first priming dose of ChAdOx1 or BNT162b2 were weakly immunogenic, raising concerns on the protection offered by such schedules. This finding indicates that not all heterologous schedules improve immunogenicity, and the immunogenicity is somewhat dependent on the sequence of certain vaccines. Further investigations of such schedules are warranted particularly for countries that have access to only CoronaVac and ChAdOx1 and are considering a mix-and-match approach.

Homologous primary series of CoronaVac, ChAdOx1 or BNT162b2 have reduced efficacy against symptomatic infection caused by SARS-CoV-2 variants, particularly the Delta^19–21^. The clinical efficacy of heterologous schedules in this study is unknown, however protection is likely to be similar or greater than homologous schedules given the association between humoral responses and vaccine efficacy^9,22^. Indeed, a recent observational study on the immunogenicity and efficacy of ChAdOx1–BNT162b2 vaccination found that this schedule provided better protection against SARS-COV-2 infection, including the variants (Omicron not tested) than the homologous BNT162b2^18^.

In LMICs that have access to COVID-19 vaccines, access to mRNA vaccines are limited. Heterologous primary series involving CoronaVac and ChAdOx1 if proven to provide similar or better protection against SARS-CoV-2 than homologous CoronaVac or ChAdOx1 series will allow more flexible COVID-19 vaccination arrangement, and potentially improve vaccine uptake. With improved vaccination coverage, particularly in LMICs, this would reduce the chance of new variants emerging and improve global equity. Evaluation of such schedule in a Thailand have revealed similar vaccine effectiveness between heterologous CoronaVac-ChAdOx1 schedule (74%) compared to homologous 2-dose ChAdOx1 (83%), and higher than 2-dose CoronaVac (60%) schedule^23^. In countries where mRNA vaccines are in limited supply, their use as a second dose in the primary series or third dose would have the benefit of providing robust immunogenicity while also potentially reducing the risks of the rare side effects such as pericarditis and myocarditis which are mainly reported after the second dose of mRNA vaccines^24,25^. In addition, repeat dosing of the same viral-vector vaccine may also induce anti-vector immunity, potentially reducing the protective immune responses which can be overcome by heterologous schedule.

It is increasingly evident that three doses of COVID-19 vaccines with homologous mRNA schedule or heterologous schedule with mRNA as booster are needed to protect against the Omicron variant^26,27^. This is in line with recent immunological studies that showed that three doses of homologous mRNA schedule or heterologous schedule with mRNA as booster induced high levels of neutralising antibodies against Delta and Omicron variants^28–30^. Consistent with this finding, we found that a third dose of BNT162b2 given to participants who received CoronaVac and ChAdOx1 primary series, in either sequence, induced high levels of neutralising antibodies against the Delta and Omicron variants. This is of interest that despite the poorer immunogenicity of ChAdOx1-CoronaVac primary series compared to when BNT162b2 are given as second dose, the BNT162b2 given as a third dose was able to induce high level humoral immunity against Omicron. This is the first study to demonstrate that neutralising antibodies against SARS-CoV-2 variants following heterologous triple platform vaccination supporting the mix-and-match principle in primary series and booster using BNT162b2 as the final dose.

Limitations of this study were that the participants were not blinded for the study vaccine, which may have influenced the reporting of adverse reactions. Secondly, the small sample size in each group did not allow us to identify any potential rare AEs. However, findings from this study have been used to inform the Thailand National COVID-19 vaccination program recommendation by including the CoronaVac-ChAdOx1 regimen during the time when mRNA vaccines were not available. Ongoing surveillance of any rare AEs and the clinical effectiveness of this heterologous schedule will inform the feasibility of such schedule against the SARS-CoV-2 variants. Lastly, the BNT162b2 booster in this study was only given to two groups who received CoronaVac and ChAdOx1, and there was no homologous three-dose BNT162b2 group for comparison of their immunogenicity. Despite this, it was clear that BNT162b2 as the third booster vaccination provided high neutralizing activity against Delta and Omicron, which is likely to provide protection against these two predominant variants. Future studies incorporating mRNA booster to other heterologous primary schedules will be of interest, particularly with other COVID-19 vaccines that have recently received WHO EUL.

In conclusion, we found that heterologous COVID-19 primary series using either ChAdOx1 or BNT162b2 as second dose were highly immunogenic against SARS-CoV-2 ancestral strain, Beta and Delta variants, but low neutralizing antibody against Omicron. A third dose of BNT162b2 given to individuals who received heterologous primary series of CoronaVac and ChAdOx1 induced high neutralising antibodies against Delta and Omicron. Larger studies including longer follow up are needed to validate these findings clinically and evaluate the persistence of immune responses following heterologous COVID-19 vaccination. Our findings have implications for countries where have implemented heterologous COVID-19 primary series involving CoronaVac and ChAdOx1, and in settings where there are critical vaccine supply issues, particularly mRNA vaccine.

## Method

### Study design

This was a single-centre prospective cohort study conducted at the Clinical Research Center of the Faculty of Medicine Siriraj Hospital (SICRES), Bangkok, Thailand. Participants were openly assigned to one of six heterologous combinations of CoronaVac, ChAdOx1 and BNT162b2 groups or a homologous BNT162b2 (as control) group. For heterologous schedules, the interval between the two doses was 4 weeks. Adverse events following the second dose were recorded to determine the safety and reactogenicity. Blood samples were collected at baseline (pre-vaccination), 4 weeks after the first dose (prime), 2 weeks and 10-12 weeks following the second dose (boost) to determine immunogenicity. The clinical study was approved by the Siriraj Institutional Ethics Review Board (approval no.Si537/2021). The procedures in this study adhere to the tenets of the Declaration of Helsinki. The study was registered at http://www.thaiclinicaltrials.org/show/TCTR20210720007.

### Study participants

Participants were healthy adults aged 18-60 years who were not known to be infected with SARS-CoV-2 or received COVID-19 vaccine other than as assigned in the study. The exclusion criteria were unstable underlying disease, having acute illness, had a history of anaphylaxis; were pregnant females, were immunocompromised or receiving immunosuppressants at screening. Participants who had a positive anti-nucleoprotein (anti-NP) or anti-RBD IgG antibody at baseline were excluded.

### Procedures

Participants were recruited into the study following informed consent and were randomised to one of seven prime-boost groups: CoronaVac-ChAdOx1, CoronaVac-BNT162b2, ChAdOx1-CoronaVac, ChAdOx1-BNT162b2, BNT162b2-CoronaVac, BNT162b2-ChAdOx1 or homologous BNT162b2. As mRNA booster vaccination as the third dose has been included in the national COVID-10 vaccination program in Thailand, we further explore the safety and immunogenicity of heterologous BNT162b2 boosting in this cohort. The participants in the CoronaVac-ChAdOx1 and ChAdOx1-CoronaVac groups had low antibody response based on preliminary analysis and were invited back to receive a BNT162b2 booster dose at 10-12 weeks after the second dose.

Following each vaccination, all participants were observed for any immediate reaction for at least 30 min. Participants were instructed to record self-assessments in an electronic diary (eDiary) for seven days after the second dose to track adverse events (AEs), which included solicited local and systemic adverse reactions. Adverse events in this study were defined as any unfavourable or unintended signs, symptoms, or disease that occurs in any participant. Solicited local AEs included pain, erythema, and swelling/induration at the injection site, and localized axillary swelling or tenderness ipsilateral to the injection arm. Solicited systemic AEs include headache, fatigue, myalgia, arthralgia, nausea/vomiting, rash, fever, and chills. The severity of solicited AEs were graded using a numerical scale from 1 to 4 based on the Toxicity Grading Scale for Healthy Adult and Adolescent Volunteers Enrolled in Preventive Vaccine Clinical Trials from the United States Food and Drug Administration (FDA)^31^.

Blood samples collected were tested for anti-receptor binding domain of SARS-CoV-2 spike protein IgG (Anti-RBD IgG) at all timepoints and T-cell response using interferon-gamma release assay (IGRA) at four weeks after the first dose and two weeks after the second dose. A qualitative anti-NP IgG was tested at baseline. The standard 50% plaque reduction neutralizing test (PRNT) assay and pseudovirion-based neutralizing test pseudotype-based neutralization assays (PVNT) were used to measure neutralizaing antibodies against SARS-CoV-2 variants on blood samples collected at two weeks after the second dose and third dose.

### Laboratory assays

The anti-RBD IgG was measured by chemiluminescent microparticle immunoassay using the SARS-CoV-2 IgG II Quant (Abbott, List No. 06S60) on the ARCHITECT i System. This assay linearly measures the level of antibody between 21.0 - 40,000.0 arbitrary unit (AU)/mL, which was converted later to WHO International Standard concentration as binding antibody unit per mL (BAU/mL) following the equation provided by the manufacturer (BAU/mL =0.142 x AU/mL). A level greater or equal to the cutoff value of 50 AU/mL or 7.1 BAU/mL was defined as seropositive. The qualitative anti-NP IgG was also measured by CMIA using the SARS-CoV-2 IgG (Abbott, List No. 06R86) on the ARCHITECT i System.

The SARS-CoV-2 live-virus neutralisation assay (PRNT_50_) was performed at the Department of Medical Science, Ministry of Public Health, Bangkok, Thailand. Briefly, the Vero cells were seeded at 2× 10^5^ cells/well/ 3 ml and placed in 37°C, 5% CO_2_ incubator for 1 day. Test sera were initially diluted at 1:10, 1:40, 1:160 and 1:640, respectively. SARS-CoV-2 virus (Delta and Beta variant) was diluted in culture medium to yield 40-120 plaques/well in the virus control wells. Cell control wells, convalescent patient serum and normal human serum were also included as assay controls. Equal volume of diluted serum and the optimal plaque numbers of SARS CoV-2 virus were incubated at 37°C in water bath for 1 hr. After removing the culture medium from Vero cell culture plates, 200 ul of the virus-serum antibody mixture were inoculated into monolayer Vero cells. The culture plates were rocked every 15 min for 1h. Three ml of overlay semisolid medium (Sigma, USA) and 10% FBS) were replaced after removing excessive viruses. All plates were incubated at 37°C, 5% CO_2_ for 7 days. Cells were fixed with 10% (v/v) formaldehyde then stained with 0.5% crystal violet in PBS. The number of plaques formed was counted in triplicate wells and percentage of plaque reduction at 50% was calculated. The PRNT_50_ of test sample was defined as the reciprocal of the highest test serum dilution for which the virus infectivity is reduced by 50% when compared with the average plaque counts of the virus control and was calculated by using a four-point linear regression method. Plaque counts for all serial dilutions of serum were scored to ensure that there was a dose response. The detection limit of PRNT_50_ was 1:10.

SARS-CoV-2 pseudovirus was constructed for pseudotype-based neutralization assays (PVNT). Codon-optimized gene encoding the spike of Omicron (B.1.1.529/ BA.1) and Delta (B.1.617.2) were generated by gene synthesis (Genscript) and cloned into the pCAGGS expressing plasmid by In-Fusion assembly (Clontech). Pseudovirus was generated and concentrated as previously described (1). Briefly, Human embryonic kidney (HEK) 293T/17 cells were transfected with the pCAGGS-S expression vector (Delta or Omicron) in conjunction with p8.9171 and pCSFLW72 using polyethylenimine (PEI, Polysciences, Warrington, USA). HIV (SARS-CoV-2) pseudotypes containing supernatants were harvested 72 hours post-transfection, aliquoted and frozen at −80°C prior to use.

PVNT were carried out as described previously^32^. Briefly, HEK293T cells overexpressing human ACE2 were maintained in Dulbecco’s modified Eagle’s medium (DMEM) supplemented with 10% FBS, 200mM L-glutamine, 100μg/ml streptomycin and 100 IU/ml penicillin. At 24 hours before the assay, cells (5×10^5^ cells/ml cells in 10 mm dish) were transfected with pCAGGS expressing codon-optimized human TMPRSS2 using Fugene HD transfection reagent (Promega). Serum samples were heat-inactivated at 56°C for 30 min and subsequently serially diluted from 1:40 to 1:5,120 in complete DMEM prior to incubation with specified SARS-CoV-2 pseudovirus, incubated for 1 h at 37°C, and plated onto TMPRSS2-expressing HEK293T-ACE2 target cells (1 × 10^4^ cells/well). After 48 h, luciferase activity was quantified by the addition of Bright-GloTM luciferase substrate (Promega) and analysis on Synergy(tm) HTX Multi-Mode Microplate Reader (BioTek). Antibody titer was then calculated by interpolating the point at which infectivity had been reduced to 50% of the value for the no serum control samples using GraphPad Prism 9.0 software.

IGRA were performed using the Quantiferon SARS-CoV-2 assay (QIAGEN) according to the manufacturer’s instruction. Fresh whole blood samples were collected into tubes containing SARS-CoV-2 S peptide pools for CD4^+^ T cells (Ag 1), and CD8^+^ T cells (Ag2), a mitogen tube as the positive control and an unstimulated tube as negative control. The specimen was incubated at 37 ° C for 16-24 hours and centrifuged to separate plasma. Interferon-gamma (IFN-γ) concentration in the plasma fraction was measured with an automated QuantiFERON SARS-CoV-2 ELISA instrument and reported in International Units per mL (IU/mL)^33,34^. The cut-off points for positivity were determined as the level above the mean plus three standard deviations of the negative control. Using 61 negative controls at the study site, the cut-offs for Ag1 and Ag2 were greater than 0.12 IU/mL and greater than 0.17 IU/mL, respectively. A positive response to either Ag1 or Ag2 was considered positive.

### Statistical Analysis

The AE endpoints were presented as frequencies (%) and compared using Fisher’s exact test. The anti-RBD IgG antibody responses were presented as geometric mean concentrations (GMC) with 95% confidence intervals (CI). PRNT_50_ and PVNT_50_ data were presented as geometric mean titers (GMT) with 95% CIs. The seroconversion rates, anti-RBD IgG GMCs, and neutralising antibody GMTs were compared within group and between the groups using paired *t* test and unpaired *t* test. The homologous two-dose CoronaVac and ChAdOx1 primary series previously reported^14^ were presented as referenced data. All statistical analyses were conducted using STATA version 17 (StataCorp, LP, College Station, TX, USA).

## Data Availability

The datasets generated during and/or analyzed during the current study are available from the corresponding author on reasonable request.

## Acknowledgement

The authors gratefully acknowledge the Siriraj Institute of Clinical Research (SICRES) team for supporting the study process and all participants who took part in this study.

## Author Contributions

Conceptualization and Methodology, S.N., P.W., K.S., S.S., Y.J., N.M., and K.C.; Formal analysis and data curation: S.N., L.J., Z.Q.T., P.V.; Project administration, S.N., K.C.; Supervision, K.C.; Resources and Funding, K.C. All involved with Investigation, and Writing-review and editing.

## Figure legends

**Supplementary Fig. S1 Correlation between plaque-reduction neutralization test (PRNT50) and pseudovirus-based neutralization test (PVNT50) against Delta variant. r= 0.78. p<0.0001**

**Supplementary Fig. S2 Interferon-gamma (IFN-γ) responses at 4 weeks and 2 weeks after first and second vaccine dose, respectively**. Interferon-gamma (IFN-γ) concentration were measured in blood after stimulation with SARS-CoV-2 peptide pool for CD4+ T cells (Ag 1) (A), and CD8+ T cells (Ag2) (B). Data presented as International Units per mL (IU/mL).

## References

1. Worldometer. COVID-19 Coronavirus Pandemic. https://www.worldometers.info/coronavirus/ (2022). accessed on 22nd February 2022

2. The New York Times. Tracking Coronavirus Vaccinations Around the World. https://www.nytimes.com/interactive/2021/world/covid-vaccinations-tracker.html (2022). accessed on 22nd February 2022

3. Mallapaty, S. et al. How COVID vaccines shaped 2021 in eight powerful charts. Nature 600, 580–583 (2021).

4. Liu, Q, et al. Effectiveness and safety of SARS-CoV-2 vaccine in real-world studies: a systematic review and meta-analysis. Infect. Dis. Poverty 10, 132 (2022).

5. Garcia-Beltran, W. F. et al. mRNA-based COVID-19 vaccine boosters induce neutralizing immunity against SARS-CoV-2 Omicron variant. Cell 185, 457–466 (2022).

6. Pajon, R. et al. SARS-CoV-2 Omicron Variant Neutralization after mRNA-1273 Booster Vaccination. N. Engl. J. Med. Online ahead of print. (2022).

7. Angkasekwinai, N. et al. The immunogenicity and safety of different COVID-19 booster vaccination following CoronaVac or ChAdOx1 nCoV-19 primary series. medRxiv preprint doi: https://doi.org/10.1101/2021.11.29.21266947 (2022).

8. Barda, N. et al. Effectiveness of a third dose of the BNT162b2 mRNA COVID-19 vaccine for preventing severe outcomes in Israel: an observational study. Lancet 398, 2093–2100 (2021).

9. Khoury, D. S. et al. Neutralizing antibody levels are highly predictive of immune protection from symptomatic SARS-CoV-2 infection. Nat. Med. 27, 1205–1211 (2021).

10. ONE. The astoundingly unequal vaccine rollout. https://www.one.org/africa/issues/covid-19-tracker/explore-vaccines/ (2022). accessed on 25th February 2022.

11. Liu, X. et al. Safety and immunogenicity of heterologous versus homologous primeboost schedules with an adenoviral vectored and mRNA COVID-19 vaccine (Com-COV): a single-blind, randomised, non-inferiority trial. Lancet 398, 856–869 (2021).

12. Schmidt, T. et al. Immunogenicity and reactogenicity of heterologous ChAdOx1 nCoV-19/mRNA vaccination. Nat. Med. 27, 1530–1535 (2021).

13. Yorsaeng, R. et al. Immune response elicited from heterologous SARS-CoV-2 vaccination: Sinovac (CoronaVac) followed by AstraZeneca (Vaxzevria). medRxiv preprint doi: https://doi.org/10.1101/2021.09.01.21262955 (2021).

14. Angkasekwinai, N. et al. Safety and Immunogenicity of CoronaVac and ChAdOx1 Against the SARS-CoV-2 Circulating Variants of Concern (Alpha, Delta, Beta) in Thai Healthcare Workers. medRxiv preprint doi: https://doi.org/10.1101/2021.10.03.21264451 (2022).

15. Groß, R. et al. Heterologous ChAdOx1 nCoV-19 and BNT162b2 prime-boost vaccination elicits potent neutralizing antibody responses and T cell reactivity eBioMedicine 75, 103761 (2021).

16. Barros-Martins, J. et al. Humoral and cellular immune response against SARS-CoV-2 variants following heterologous and homologous ChAdOx1 nCoV-19/BNT162b2 vaccination. medRxiv preprint doi: https://doi.org/10.1101/2021.06.01.21258172 (2021).

17. Borobia, A. M. et al. Immunogenicity and reactogenicity of BNT162b2 booster in ChAdOx1-S-primed participants (CombiVacS): a multicentre, open-label, randomised, controlled, phase 2 trial. Lancet 398, 121–130 (2021).

18. Pozzetto, B. et al. Immunogenicity and efficacy of heterologous ChAdOx1–BNT162b2 vaccination. Nature 600, 701–706 (2021).

19. Berna, J. L. et al. Effectiveness of Covid-19 Vaccines against the B.1.617.2 (Delta) Variant. N. Engl. J. Med. 385, 585–594 (2021)

20. Vacharathit, V. et al. CoronaVac induces lower neutralising activity against variants of concern than natural infection. Lancet. Infect. Dis. 21, 1352–1354 (2021).

21. AlQahtani, M. et al. Morbidity and mortality from COVID-19 postvaccination breakthrough infections in association with vaccines and the emergence of variants in Bahrain. https://assets.researchsquare.com/files/rs-828021/v1_covered.pdf?c=1631876901. accessed on 25th February 2022.

22. Angyal, A. et al. T-cell and antibody responses to first BNT162b2 vaccine dose in previously infected and SARS-CoV-2-naive UK health-care workers: a multicentre prospective cohort study. Lancet Microbe. 3, e21–31 (2022).

23. Sritipsukho, P. et al. Comparing real-life effectiveness of various COVID-19 vaccine regimens during the delta variant-dominant pandemic: a test-negative case-control study. Emerg. Microbes. Infect. 11, 585–592 (2022).

24. Mevorach, D. et al. Myocarditis after BNT162b2 mRNA Vaccine against Covid-19 in Israel. N. Engl. J. Med. 385, 2140–2149 (2021).

25. Bozkurt, B. et al. Myocarditis With COVID-19 mRNA Vaccines. Circulation 144, 471–484 (2021).

26. Accorsi, E. K. et al. Association Between 3 Doses of mRNA COVID-19 Vaccine and Symptomatic Infection Caused by the SARS-CoV-2 Omicron and Delta Variants. JAMA 327, 639–651 (2022).

27. Ferdinands, J. M. Waning 2-Dose and 3-Dose Effectiveness of mRNA Vaccines Against COVID-19–Associated Emergency Department and Urgent Care Encounters and Hospitalizations Among Adults During Periods of Delta and Omicron Variant Predominance — VISION Network, 10 States, August 2021–January 2022. MMWR 71, 255–263 (2022).

28. Clemens, S. A. C. et al. Heterologous versus homologous COVID-19 booster vaccination in previous recipients of two doses of CoronaVac COVID-19 vaccine in Brazil (RHH-001): a phase 4, non-inferiority, single blind, randomised study. Lancet 399, 521–529 (2022).

29. Cheng, S. M. S. et al. Neutralizing antibodies against the SARS-CoV-2 Omicron variant BA.1 following homologous and heterologous CoronaVac or BNT162b2 vaccination. Nat. Med. https://doi.org/10.1038/s41591-022-01704-7 (2022).

30. Pérez-Then, E. et al. Neutralizing antibodies against the SARS-CoV-2 Delta and Omicron variants following heterologous CoronaVac plus BNT162b2 booster vaccination. Nat. Med. https://doi.org/10.1038/s41591-022-01705-6 (2022).

31. U.S. Department of Health and Human Services, Food and Drug Administration, Center for Biologics Evaluation and Research. Guidance for Industry Toxicity Grading Scale for Healthy Adult and Adolescent Volunteers Enrolled in Preventive Vaccine Clinical Trials. https://www.fda.gov/media/73679/download (2007).

32. Koonpaew, S. et al. A Single-Cycle Influenza A Virus-Based SARS-CoV-2 Vaccine Elicits Potent Immune Responses in a Mouse Model. Vaccines 9, 850 (2021).

33. Murugesan, K. et al. Interferon-γ Release Assay for Accurate Detection of Severe Acute Respiratory Syndrome Coronavirus 2 T-Cell Response. Clin. Infect. Dis. 73, e3130–e3132 (2021).

34. Martínez-Gallo, M. et al. Commercialized kit to assess T-cell responses against SARS-CoV-2 S peptides. A pilot study in Health Care Workers. medRxiv preprint https://doi.org/10.1101/2021.03.31.21254472 (2021).

