## Supplementary Table S1 for "Immunogenicity and reactogenicity against the SARS-CoV-2 variants following heterologous primary series involving CoronaVac and ChAdOx1 and BNT162b2 plus heterologous BNT162b2 booster vaccination: An open-label randomized study in healthy Thai adults"

**Supplementary Table S1. Immunogenicity of heterologous schedules of CoronaVac, ChAdOx1, BNT162b2 and homologous BNT162b2 schedule.**

|  | Type of heterologous and homologous vaccinations | | | | | | |
| --- | --- | --- | --- | --- | --- | --- | --- |
|  | CoronaVac-ChAdOx1 | CoronaVac-BNT162b2 | ChAdOx1- CoronaVac | ChAdOx1- BNT162b2 | BNT162b2-CoronaVac | BNT162b2-ChAdOx1 | BNT162b2-BNT162b2 |
| At 4 weeks after first dose vaccination | n=30 | n=30 | n=29 | n=26 | n=29 | n=30 | n=27 |
| Anti-RBD IgG, GMC (95%CI) (BAU/mL) | 15.40  (10.40, 22.81) | 19.77  (13.35, 29.29) | 83.02(59.22, 116.38) | 93.37  (63.56, 137.15) | 137.55  (91.16, 207.56) | 114.27  (84.71, 154.13) | 102.35  (74.93, 139.79) |
| IGRA Positive, n (%) | 7 (23.33) | 1 (3.33) | 17 (58.62) | 11 (42.31) | 8 (27.59) | 12 (41.38) | 7 (25.93) |
| At 2 weeks after second dose vaccination | n=30 | n=30 | n=29 | n=26 | n=29 | n=30 | n=26 |
| Anti-RBD IgG, GMC (95%CI) (BAU/mL) | 851.40 (799.05, 1318.39) | 2,181.84 (2079.94, 3870.63) | 137.04 (127.93, 221.36) | 2,132.68 (1920.87, 3,010.06) | 255.21 (205.24, 334.98) | 1,201.16 (1092.68, 1817.69) | 2,248.76 (2068.72, 3561.77) |
| PRNT_50_, GMT (95%CI) against Delta variant | 78.65  (57.09, 108.34) | 196.97  (126.91, 305.68) | 20.36  (14.18, 29.21) | 195.12  (161.46, 235.74) | 22.69  (16.00, 32.19) | 112.36  (85.82, 147.11) | 178.90  (136.67, 234.12) |
| PRNT_50_, GMT (95%CI) against Beta variant | 30.55  (18.52, 50.40) | 62.36  (38.27, 101.62) | 9.01  (7.26, 11.19) | 43.28  (28.86, 64.93) | 8.18  (6.48, 10.33) | 20.43  (14.41, 28.96) | 40.06  (25.03, 64.09) |
| PRNT_50_, GMR (95%CI) (Delta/Beta) after second vaccination) | 2.57  (1.69, 3.93) | 3.15  (2.46, 4.06) | 2.26  (1.60, 3.19) | 4.51  (3.42, 5.94) | 2.77  (1.86, 4.12) | 5.50  (4.11, 7.37) | 4.47  (3.15, 6.33) |
| PVNT_50_, GMT (95%CI) against Delta variant | 235.25 (176.77, 313.08) | 565.06 (331.35, 963.62) | 19.28  (8.66, 42.94) | 681.23 (534.10, 868.90) | 28.54  (11.76, 69.27) | 249.67 (176.07, 354.04) | 748.39  (535.60, 1045.73) |
| PVNT_50_, GMT (95%CI) against Omicron variant | 1.03  (0.39, 2.72) | 20.13  (7.46, 54.33) | 0.36  (0.15, 0.89) | 12.14  (4.30, 34.23) | 0.28  (0.13, 0.61) | 1.90  (0.62, 5.83) | 6.26  (2.15, 18.22) |
| PVNT_50_, GMR (95%CI) (Delta/Omicron) after second vaccination | 229.34  (87.27, 602.69) | 28.07  (12.15, 64.86) | 52.93  (17.55, 159.65) | 56.13  (22.63) | 102.67  (30.51, 345.49) | 131.59  (47.44, 364.95) | 119.63  (45.63, 313.62) |
| IGRA Positive, n (%) | 20 (66.67) | 23 (76.67) | 18 (62.07) | 18 (69.23) | 16 (55.7) | 22 (73.33) | 21 (80.77) |
| At 10-12 weeks after second dose vaccination | n=30 | n=30 | n=29 | n=24 | n=27 | n=30 | n=25 |
| Anti-RBD IgG, GMC (95%CI) (BAU/mL) | 223.23  (165.15, 298.13) | 460.39  (359.29, 589.92) | 44.20  (33.18, 58.89) | 506.80  (391.28, 656.43) | 62.08  (48.08, 80.15) | 301.61  (237.65, 382.77) | 488.48  (367.81, 648.75) |
| Anti-RBD IgG, GMR (95%CI) 2 weeks/10-12 weeks after second vaccination | 3.81  (3.21, 4.53) | 4.74  (3.81, 5.89) | 2.88  (2.41, 3.43) | 4.15  (3.69, 4.66) | 3.75  (3.17, 4.44) | 3.98  (3.50, 4.53) | 4.72  (4.16,5.36) |
| At 2 weeks after third dose vaccination | n=30 | NA | n=29 | NA | NA | NA | NA |
| Anti-RBD IgG, GMC (95%CI) (BAU/mL) | 2,518.81 (1960.37, 3236.35) | NA | 2,610.58 (2037.70, 3344.52) | NA | NA | NA | NA |
| Anti-RBD IgG, GMR (95%CI) 2 weeks/10-12 weeks after second vaccination | 2.46  (1.93, 3.13) | NA | 18.93  (14.93, 24.00) | NA | NA | NA | NA |
| PVNT_50_: GMT (95%CI) against Delta variant | 528.42  (340.96, 818.95) | NA | 1,085.69 (790.28, 1491.52) | NA | NA | NA | NA |
| PVNT_50_ Delta, GMR (95% CI) post 3^rd^ dose/post 2^nd^ dose | 2.25  (1.54, 3.28) | NA | 70.89  (30.29, 165.93) | NA | NA | NA | NA |
| PVNT_50_, GMT (95%CI) against Omicron variant | 149.51  (71.87, 311.02) | NA | 419.14 (316.16, 555.66) | NA | NA | NA | NA |
| PVNT50 Omicron, GMR (95% CI) post 3^rd^ dose/post 2^nd^ dose | 145.75  (50.27, 422.63) | NA | 1,150.55 (464.16, 2851.93) | NA | NA | NA | NA |
| PVNT_50_, GMR (95% CI) post 3^rd^ dose (Delta/Omicron) | 3.53  (1.89, 6.59) | NA | 2.59  (2.08, 3.22) | NA | NA | NA | NA |
