## Supplementary figures and images for "Immunogenicity and reactogenicity against the SARS-CoV-2 variants following heterologous primary series involving CoronaVac and ChAdOx1 and BNT162b2 plus heterologous BNT162b2 booster vaccination: An open-label randomized study in healthy Thai adults"

### Supplementary Figure S1

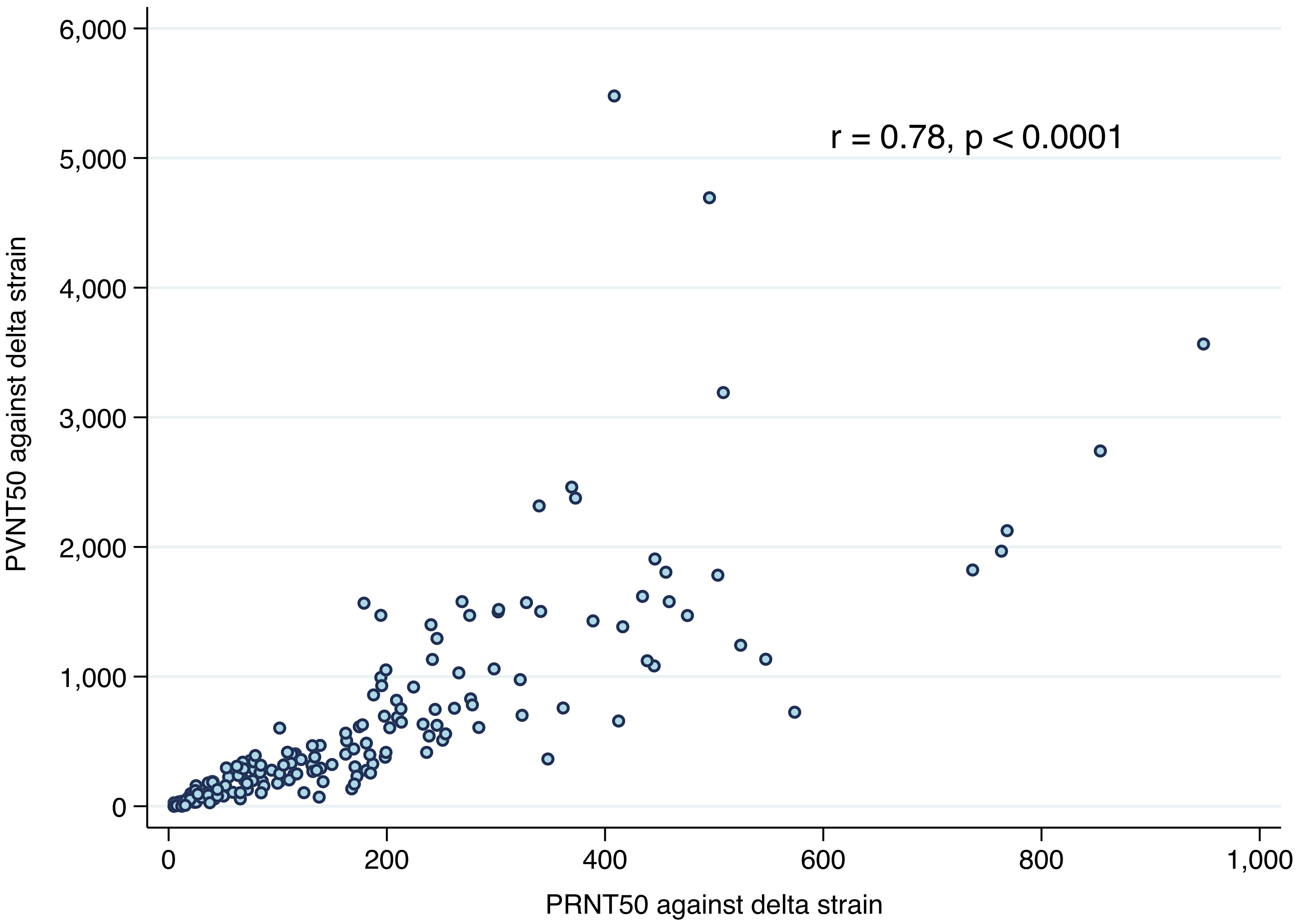

### Supplementary Figure S1

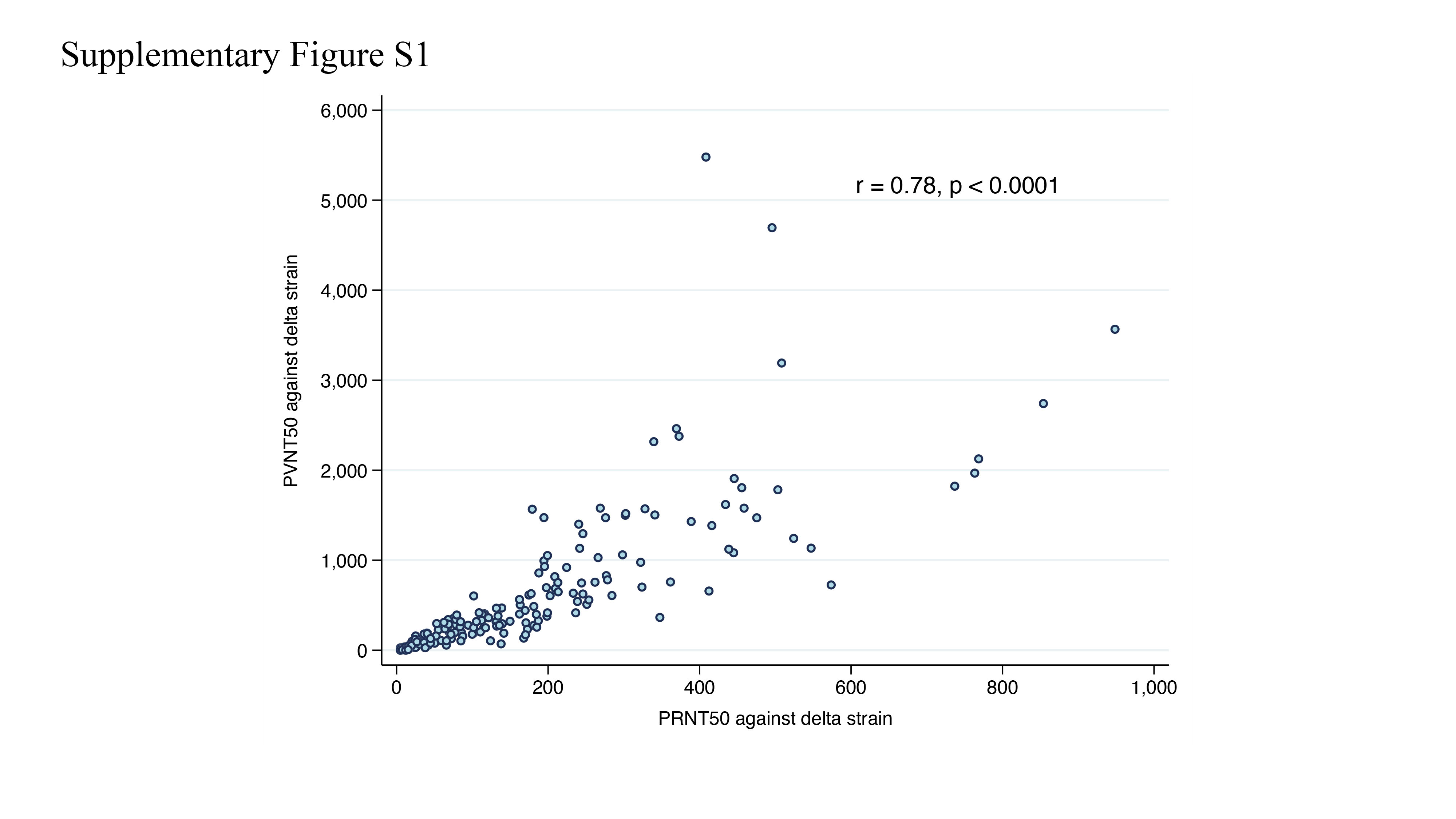

### Supplementary Figure S2

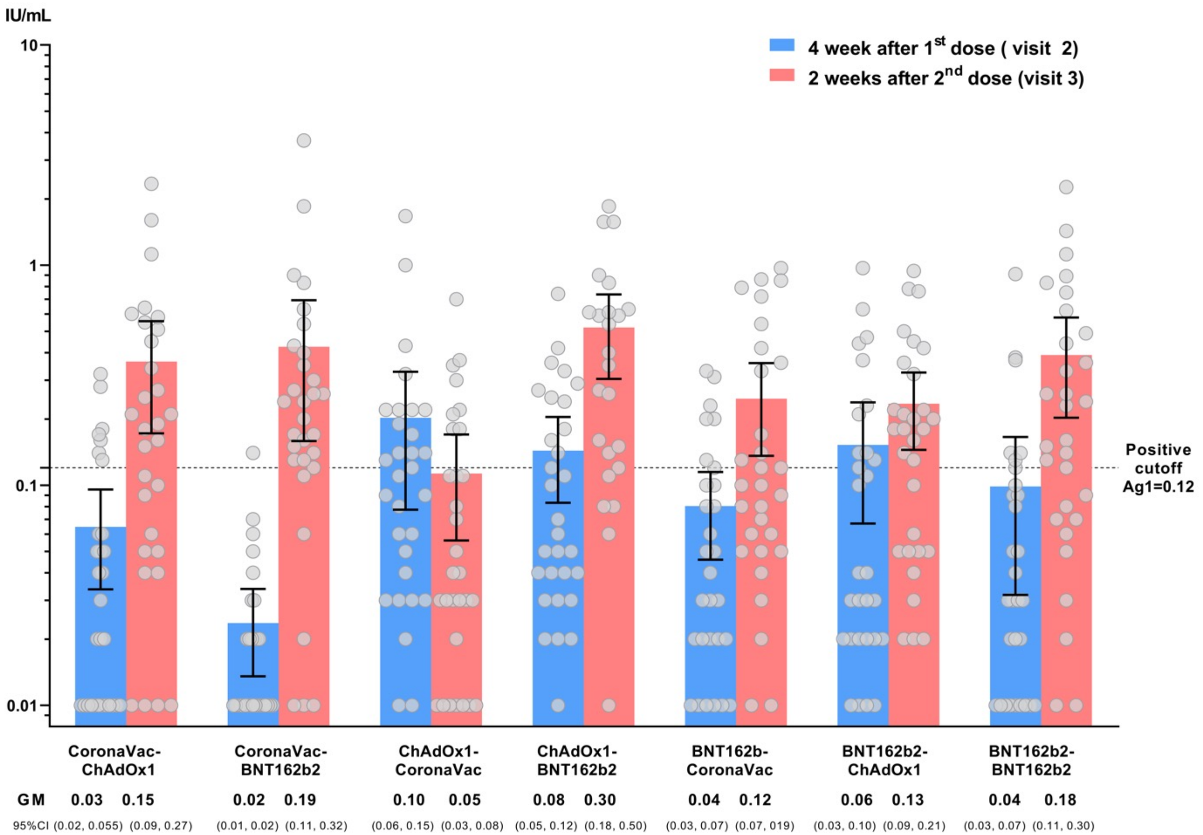

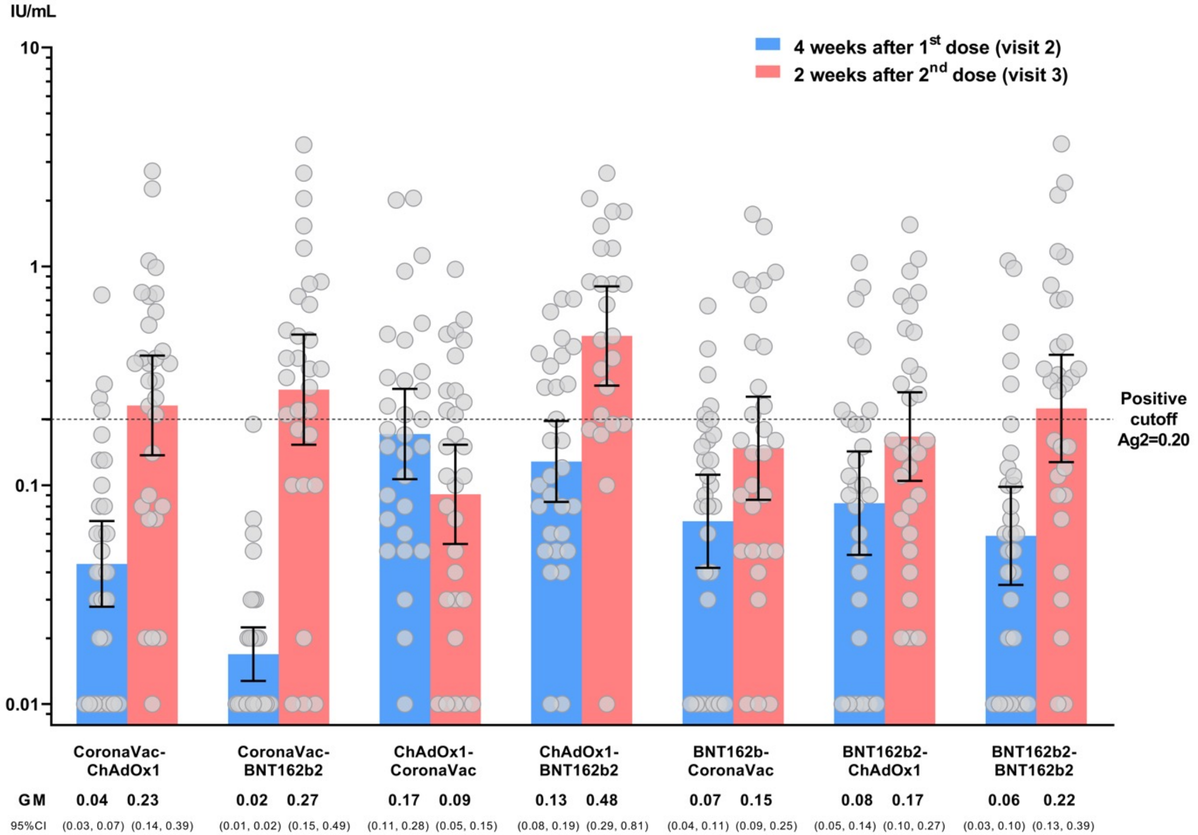

### Supplementary Figure S2

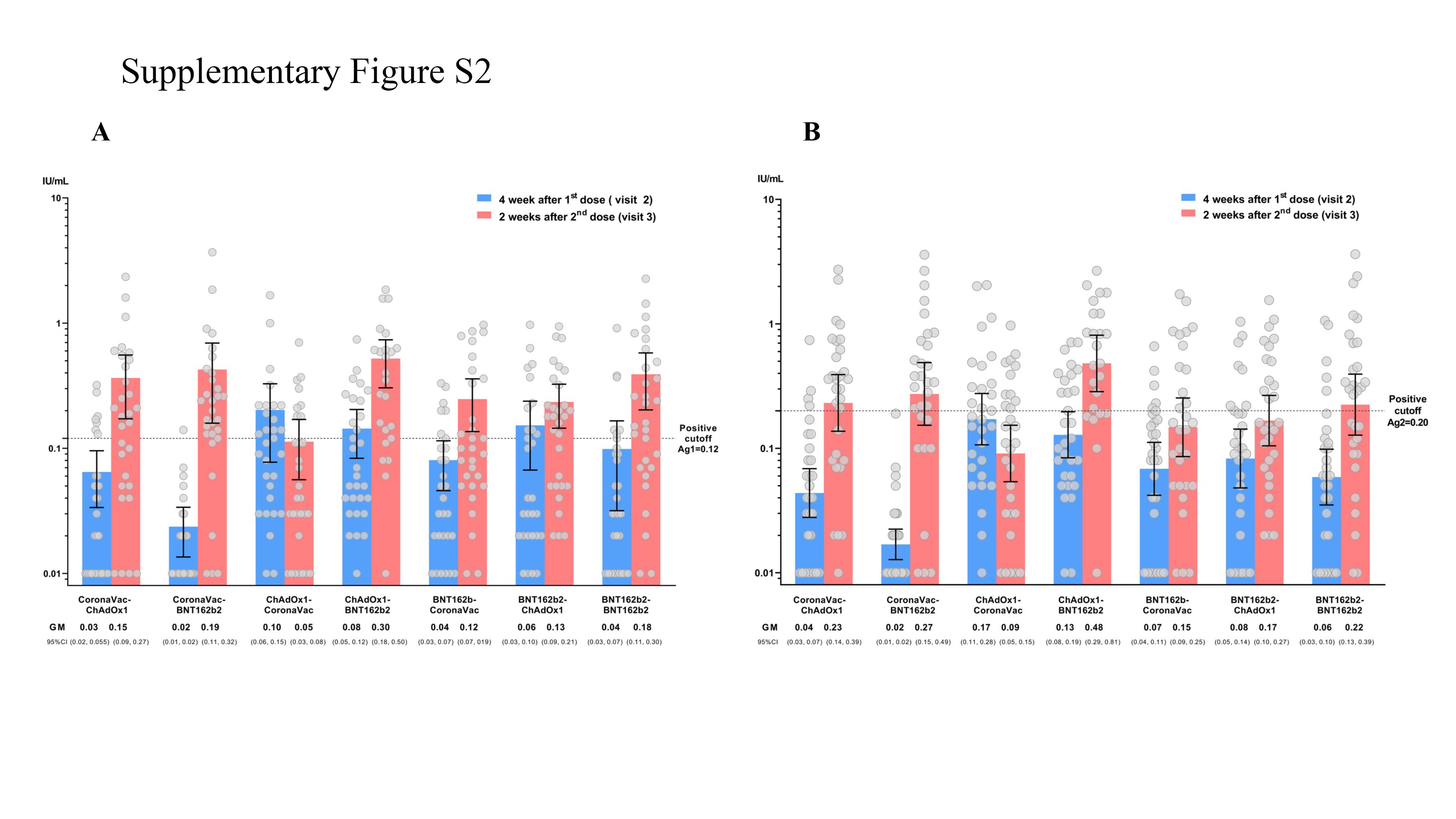
